# Sleep architecture correlates with neurological and neurobehavioral short- and mid-term outcome in a sample of very preterm infants

**DOI:** 10.1101/2024.12.13.24318989

**Authors:** Sara Uccella, Valentina Marazzotta, Deborah Preiti, Marcella Battaglini, Gaia Burlando, Monica Roascio, Andrea Rossi, Gabriele Arnulfo, Luca Antonio Ramenghi, Lino Nobili

**Affiliations:** Università degli Studi di Genova, Dipartimento di Neuroscienze, Riabilitazione, Oftalmologia, Genetica e Scienze Materno-Infantili DINOGMI, Genova Italia; Unità di Neuropsichiatria Infantile, Istituto Giannina Gaslini, Genova Italia; Unità di Psicologia Clinica, Istituto Giannina Gaslini, Genova Italia; Unità di Patologia Neonatale, Istituto Giannina Gaslini, Genova Italia; Università degli Studi di Genova, Dipartimento di informatica, bioingegneria, robotica e ingegneria dei sistemi, DIBRIS, Genova Italia; Unità Neuroradiologia, Istituto Giannina Gaslini, Genova Italia

**Keywords:** sleep, prematurity, sleep architecture, neurodevelopmental outcome, sleep ontogenesis

## Abstract

Newborns spend most of their time sleeping. This activity fosters neurodevelopment. Prematurity, defined by birth occurring prior the 37^th^ week of gestation, disrupts normal brain *in-utero* programming, with long-lasting consequences that carry a high social burden. Sleep alterations may contribute to these sequelae. In this pilot study we aimed to describe the 24-hours distribution of sleep states among very preterm infants (VPI), and to correlate it with neurobehavioral assessment up to 6 months of corrected age (CA). Secondly, we aimed to assess if the presence of a brain lesion detected at MRI could affect sleep duration, architecture, and quality.

Ten VPI were assessed at 32 weeks PMA with a 24-hours video-polysomnography and received a neurobehavioral examination at the time of the recording, at term equivalent age (TEA), and at 6 months CA. Analysis of sleep stages distribution and transitions, and power spectra were conducted.

Total sleep time and amount of quiet sleep positively correlated with neurological, and neurobehavioral assessment at 32 weeks PMA, at TEA, and at 6 months CA, while Sleep Onset Active Sleep (SOAS) had a negative association. Infants carrying brain lesions showed lower Total Sleep time accompanied by a higher prevalence of AS+SOAS and showed a gradient for higher power of posterior slow activity (slow δ and δ) during SOAS in the left hemisphere posterior regions.

Understanding sleep dynamics among preterm infants might provide future therapeutic/management strategies, which need to encompass sleep care.

## Introduction

Sleep is a vital, reversible state essential for health (Carskadon, 2011), regulated by homeostatic pressure and circadian rhythms(Frank, 2020). Both Non-Rapid Eye Movement (NREM) and Rapid Eye Movement (REM) sleep are crucial for neuronal plasticity and memory formation (Diekelmann & Born, 2010). NREM sleep supports the effective transferring of information between the hippocampus and cortex (Khodagholy et al., 2017) and controls synaptic downscaling (de Vivo et al., 2017). REM sleep is key for learning(Li et al., 2017), brain development, adaptive memory, and behavior(Frank & Heller, 2019) through synaptic pruning and strengthening (Aime et al., 2022).

Sleep undergoes changes from foetal to adult life, with specific patterns typical for age, associated with the development, maturation, and connectivity within neural networks (Cao et al., 2020; Dereymaeker et al., 2017). Newborns and infants spend most of their time sleeping a believed immature REM sleep, conventionally named Active Sleep (AS) because of the presence of movements, alternated with the calmer behavioural sleep state of Quiet Sleep (QS), thought to be related to adult NREM sleep (Grigg-Damberger, 2016).

One over 10 neonates are born before 37 weeks of gestation (World Health Organization, 2024). The destruction of the fetoplacental unit caused by premature birth alters normal *in-utero* sleep processes (Bennet et al., 2018), leading to future neurodevelopmental problems and psychiatric disorders, even in absence of major brain lesions (Kroll et al., 2018; Malova et al., 2023; Potenzieri et al., 2024; Uccella et al., 2023).

The sleep architecture of preterm infants remains largely unexplored (Calciolari & Montirosso, 2011; Cirelli & Tononi, 2015; Gogou et al., 2019), with limited evidence of differences between AS and QS in the most premature subjects (very preterm infants, VPI) when compared to term babies (Tokariev et al., 2019). Moreover, information regarding VPI sleep state organization and their association with future outcomes is lacking (Biagioni et al., 2005; Curzi-Dascalova et al., 1993). Transitional states are also poorly investigated but can be sources of information for understanding the neurodevelopmental trajectories of these babies (Uchitel et al., 2022).

Disruption of sleep architecture and chronic sleep loss during the neonatal period and infancy could hinder the proper formation of mature neural circuits, potentially leading to neurodevelopmental and behavioural issues (Bellesi et al., 2018). Prematurity, especially the most extreme one, can primarily affect sleep, but also the environmental stressors within Neonatal Intensive Care Unit (NICU) can be detrimental for preterms’ sleep maintenance. However, the short- and long-term impacts on abnormal neurodevelopment of sleep alterations caused by prematurity remain poorly understood.

For the first time, in this study we aim to describe sleep architecture and sleep transitions in VPI with and without brain lesions and correlate it with later neurological development. We observed VPI preterm infants during 24-hours video-polysomnographic recording to correlate sleep states, sleep stability, and dynamics of neuronal oscillations with neurobehavioral assessment at 32 weeks of post-menstrual age (PMA), at term equivalent age (TEA), and at 6 months of corrected age (CA).

## Materials and methods

This is a prospective cross-sectional study. Prior to starting this study, Ethical Committee Approval has been achieved (CERL number approval 0028224/21 of the 6/10/2021), and parents provided informed consent for the study.

### Subjects

VPI subjects born very preterm (born before the 32 weeks of PMA) and hospitalized at the NICU of the Gaslini Children Hospital, Genova (Italy) from November 2021 to May 2022 were considered eligible for the study. The ones carrying known or discovered genetic/metabolic and malformative disorders were excluded.

Patients started the protocol once they became hemodynamically and respiratory stable and free of sedations (such as morphine, barbiturates, and benzodiazepines) at 32 weeks of PMA.

Prenatal, intrapartum, and neonatal data were collected after the scoring of the neurophysiological and neurobehavioral assessment following previous studies (Uccella et al., 2023). Presence of anatomically detectable acquired brain lesions at brain MRI performed at TEA during spontaneous sleep using the “feed and wrap” technique (Ibrahim et al., 2015) on a 3T MR scanner (InteraAchieva 2.6; Philips, Best, The Netherlands) using a dedicated paediatric head/spine coil was noted. Type of acquired brain lesions (which are well-known risk factors for adverse neurodevelopment) was categorized as reported in previous studies (Malova et al., 2023,; Uccella et al., 2023).

### Sleep assessment

Video-polysmonographic assessments were obtained with a Brain QUICK^®^ (Micromed) system. Registrations were conducted for 24 consecutive hours starting at 8.00 am. Ten scalp silver electrodes were placed according the International 10-20 system (Fp1, Fp2, C3, C4, T3, T4 O1, O2, a physical reference and a ground electrode), accompanied by two electroculograms, the submental electromyogram, the cardiac and the abdominal breath electrodes, two electromyograms placed at the deltoids and a saturimetry probe (André et al., 2010; Grigg-Damberger, 2016). Electroencephalogram activity sampling frequency was set at 512 Hz.

Electrode assembly was performed in respect of individualized care measures appropriate for age (Stjerna et al., 2012). Recordings were acquired and then manually scored by the same clinician (SU), blinded for the medical history of the patient, in 30-seconds epochs, as proposed by the American Academy of Sleep Medicine (Grigg-Damberger, 2016), in order to determine Active/Quiet Wakefulness (AW and QW), AS, QS, or Transitional Sleep (TS).

AS was defined by eyes closed, intermittent eyes movements, brief facial and body movements (twitches), isolated vocalizations, variable/augmented heart rate and breathing, and continuous EEG, with mylohyoid tone absent or briefly present (*Figure 1a*). Sleep Onset Active Sleep (SOAS) was usually scored if the baby fell asleep during AS (*Figure 1b*). Mixed pattern is often seen during SOAS, low voltage irregular and, sometimes, high voltage slows patterns are frequently seen during AS (André et al., 2010; Grigg-Damberger, 2016). QS was defined by eyes closed, poor movements, deep breathing and regular heart rate, presence of mylohyoid tone, and discontinuous EEG pattern (*tracé alternant*) (*Figure 1c*). TS was scored when polysomnographic patterns were equally attributable at both SOAS+AS and QS (Grigg-Damberger, 2016) (*Figure 1d*).

**Figure 1.**
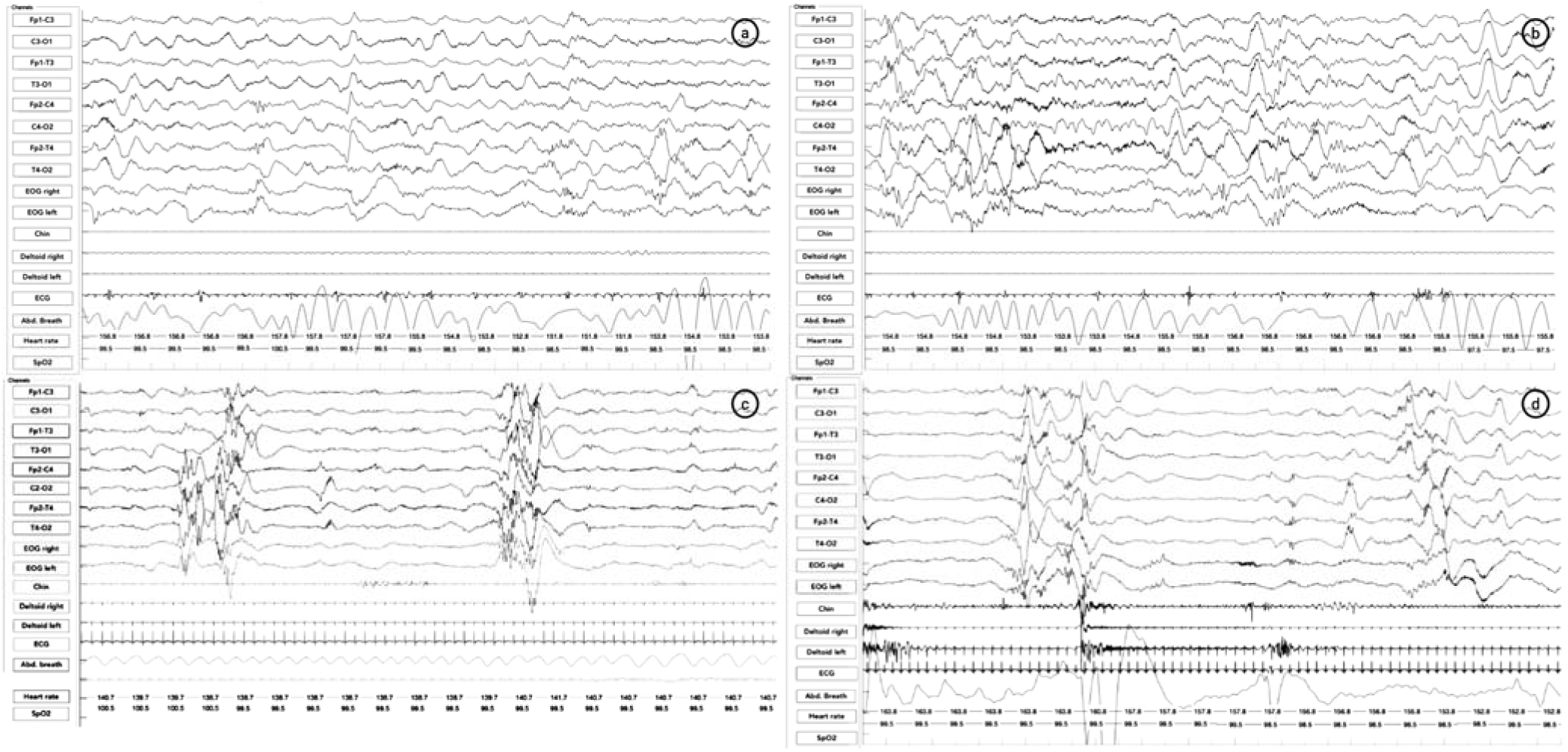
Examples of polysomnographic behavioural states recorded at 32 weeks PMA. Example of behavioural states in a very preterm infant recorded with polysomnography at 32.5 weeks PMA. *a)* Example of active sleep (AS), Note the continuous activity, the irregular breath and heart rate. *b)* Example of sleep onset active sleep (SOAS). Note the continuous activity, the irregular breath and heart rate. SOAS usually is believed to have also more REMs, present in this page. Note on 3b a muscular twitch of the cheeks. *c)* Example of quiet sleep (QS). Note the discontinuity of the traces and the alternance of burst of high amplitude waves with low-voltage highly sporadic activity in presence of chin tone, regular breath, and heart rate small variability. *d)* Example of transitional sleep. Note the discontinuos EEG pattern associated to chin tone suggest QS stage, but irregular breath and heart rate and muscle activation (deltoids), and some eye movements are in favour of AS.

Acquired polysomnographic data were transformed to EDF (European Data Format) to be imported into MatLab (MatLab 9.10, The Matworks Inc., Natick, MA, USA). Scoring data were then performed and stored in a software (PSG Lab) linked to the traces, developed in the laboratory of Institute of Molecular Bioimaging and Physiology, CNR, Genoa, Italy, which facilitates quantitative EEG analysis (Nobili et al., 2011).

### State-dependent transition probability

To further characterize the architecture of sleep in VPI, we quantified the transition probability matrix across the entire 24h recording. In the characterization of the dynamics of a system with limited possible states, the transition probably matrix describes the probability of transitioning from one state to any other states. We constructed the transition probability matrix revising the 24 hours-long hypnograms and quantifying, for a given initial state, the number of transitions towards any other state divided by the total possible transitions (*Figure 3a*). Given the instable nature of preterm sleep and the difficulties in identifying clear states, in the probability calculations we considered only the transitions to states that lasted for at least 60 seconds.

### Spectral EEG analysis

The above mentioned served for spectral EEG analysis. To estimate power spectral density, we used the Welch method. EEG signals were filtered at high-frequency filter of 70 Hz, low-frequency filter of 0.5 Hz. Artifacts (excessive movement, ventilatory machines artifacts, and instrumental noise) were removed. Inter burst low amplitude activity present in QS and TS was also removed. The bipolar Fp2-C4, T4-O2, Fp1-C3 and T3-O1 channels were chosen for the analysis to catch differences among anterior and posterior regions in the two hemispheres, in the hypothesis that differences in brain maturation, that show a posterior-anterior gradient (Brody et al., 1987; Dubois et al., 2008), could be observed. The best consecutive 10 minutes (free of artifacts) of each sleep recording were analysed. A mean spectrum was estimated for 1-minute intervals.

Mean amplitude values were obtained from each sleep stage. Five frequency bands were obtained [slow delta (δ), 0–2 Hz, delta (δ), 2–4 Hz; theta (θ), 4–8 Hz; alpha (α), 8–12 Hz; and beta (β), 12.0–24 Hz] and expressed as mV/min. Total power over the entire frequency range of interest (0.5–30 Hz) was also generated. Power spectra expressed in terms of absolute values (over total spectral power) were calculated. To analyse possible differences in brain gradient maturation, a Δ was calculated among posterior and anterior relative powers for both hemispheres for each band.

### Neurological and neurobehavioral assessment

Infants were assessed midway between feedings in a quiet environment, at the presence of their caregiver, by a trained examiner (SU) blinded to medical information at the moment of the evaluation (at 32 weeks PMA, at TEA, and at 6 months of CA). The Hammersmith Neonatal Neurological Examination (HNNE) (Dubowitz et al., 1998) and the Neonatal Behavioural Assessment Scale (NBAS) (Brazelton, 1984) were chosen for assessing their neurological and behavioural pattern at 32 weeks PMA and at TEA. Higher scores correspond to better performances. Visual performances were evaluated by visual assessment neonatal battery developed by Ricci et al. Lower scores indicate better visual performance (Ricci et al., 2008).

Neurodevelopmental assessment at 6 months of CA was performed using the Italian adaptation of the Griffiths Mental Development Scales – III edition (GMDS) (Lanfranchi et al., 2019). The Following domains were evaluated: Learning foundations, Language and Communication, Eye and Hand Coordination, Personal-Social–Emotional, and Gross Motor Skills. Scores are converted to standardized development quotients, DQ, (mean range of 100 +/- 15).

### Statistical Analysis

Statistical analyses were performed using MatLab (MatLab 9.10, The Matworks Inc., Natick, MA, USA).

The Shapiro–Wilk test was performed to evaluate the distribution of continuous variables, and outliers were checked by visual assessment of distribution graphs.

Continuous variables are reported in terms of either medians and minimum-maximum values or means and standard deviations. Categorical variables were reported in terms of absolute frequencies and percentages.

Fisher Exact or Mann-U Whitney test were considered appropriate to analyse differences among variables. Associations were calculated with Spearman correlation test. All the tests were two-sided and statistical significance was set at a *p* ≤ 0.05 with a 99% confidence interval. Borderline significance (*p* > 0.05 and ≤ 0.08) was reported. Bonferroni multiple-comparison correction was applied.

For having an exploratory view on sleep/wakefulness distribution among the sample, we divided the sleep hours in day time (08:00-20:00) and night time (20:00-08:00), in accordance with previous studies (Giganti et al., 2007). Wilcoxon rank-sum test was adopted for determining significant differences among these two times.

## Results

Ten subjects born VPI were enrolled. All variables showed a skewed non-normal distribution. *Supplementary Table 1* reports prenatal, intrapartum, and neonatal features and the prevalence of brain lesions detected at bran MRI at TEA of the sample. Moderate/severe brain lesion among the sample was 1 severe IVH (grade IV) and 1 moderate IVH (grade III), interesting more the left hemisphere. Mild brain lesions were 2 posterior punctate WML and 1 IVH grade I-II.

*Supplementary Table 2* summarises the scores obtained at the neurobehavioral assessments at 32 weeks PMA, at TEA and at 6 months of CA. Median ranges at the various developmental stages were similar to the ones observed in other samples of VPI(Canals et al., 2011; Mercuri et al., 2003; Ricci et al., 2010, 2010; Spittle et al., 2016; Wolf et al., 2002).

*Table 1* summarises polysomnographic values obtained by the 24-hours video-PSG scoring. Within the 24 hours, Mean Total Sleep time was the 70.8%, with AS being 40.1% (14.7% SOAS and 25.4 AS), QS the 27.5%, and TS the 32.4%.

**Table 1.** Distribution of behavioural stages. Data are expressed in mean minutes and percentages are referred to the whole registration (24 hours) for the Total Sleep time and to total sleep time for the different behavioural states. SOAS stands for Sleep Onset Active Sleep and AS stands for Active Sleep.

|  | *mean ± sds (%)* |
| --- | --- |
| **Total Sleep time** | **1020.8 ± 70.3 (70.8)** |
| **Total Active Sleep (SOAS+AS)** | **408.7 ± 121.8 (40.1)** |
| Sleep Onset Active Sleep | 150.4 ± 68.11 (14.7) |
| Active Sleep | 258.3 ± 90.9 (25.4) |
| **Quiet Sleep** | **280.8 ± 63.27 (27.5)** |
| **Transitional Sleep** | **331.3 ± 141.78 (32.4)** |

### VPI show different trends in sleep stages distribution during the 24 hours

In the examined sample a trend of less sleep quantity during daytime hours (08:00-20:00) than in night-time (20:00-08:00) was observed (*Figure 2*). This trend affects all types of states examined and has statistical significance for SOAS (*p* = 0.049).

**Figure 2.**
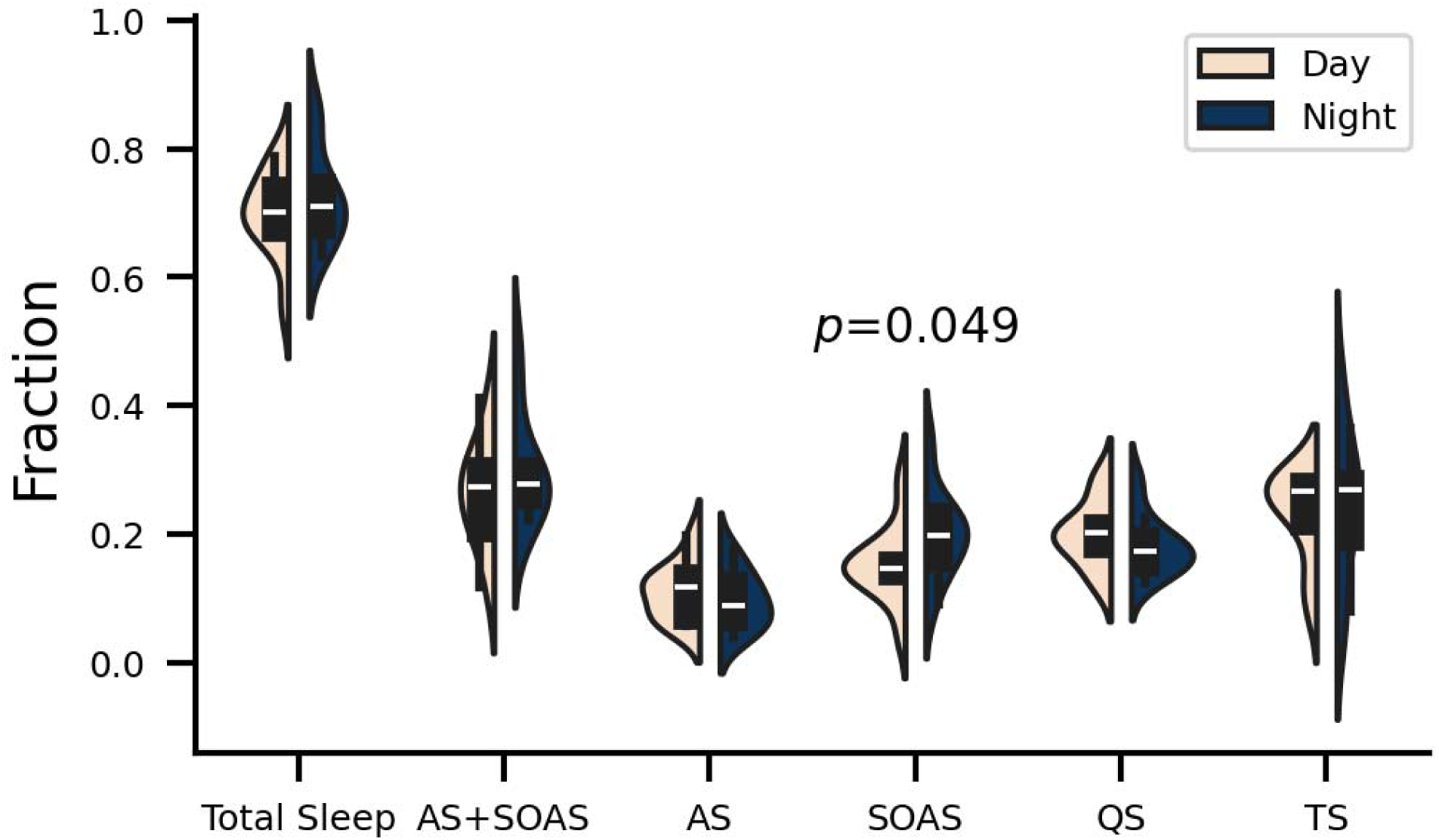
Average distribution of sleep states among day and night times. Distribution of sleep events from the total duration separate in day (light orange) and night (dark blue). Boxes show median (white ticks) and inter-quartile distances (black lines). Text label indicate p-values for significance. Only significant tests are are shown. Distributions are estimated using a kernel density estimation algorithm. AS, Active Sleep; QS, Quiet Sleep; SOAS, Sleep Onset Active Sleep; TS Transitional Sleep.

When looking at the average distribution of the hourly amount of sleep during the 24 hours, we observed a more frequent decrease of sleep time below 60% during day time (08.00-20.00) (*Supplementary* Figure 1).

### Transition probabilities differ during day or night time and are affected by presence of brain lesions

In the dynamics of preterm infants sleep, within-state transitions (diagonal elements in the transition probability matrix) were more probable than between state transitions, with QS being the most probable and hence most stable state. TS represented an unstable state with equal probability of transitioning to any other states. AW and QW together with SOAS represented a group of frequent transitional states (*Figure 3b*) with similar probability of evolving one in any others in both directions. Of interest, transition probabilities differ during day or night time. In particular, QW to AS was significantly more probable (*p*=0.036, d = −0.96) during night hours rather during daytime. SOAS to QS was significantly (*p*=0.04, d = 0.94) more probable during day time (*Figure 3c*).

**Figure 3.**
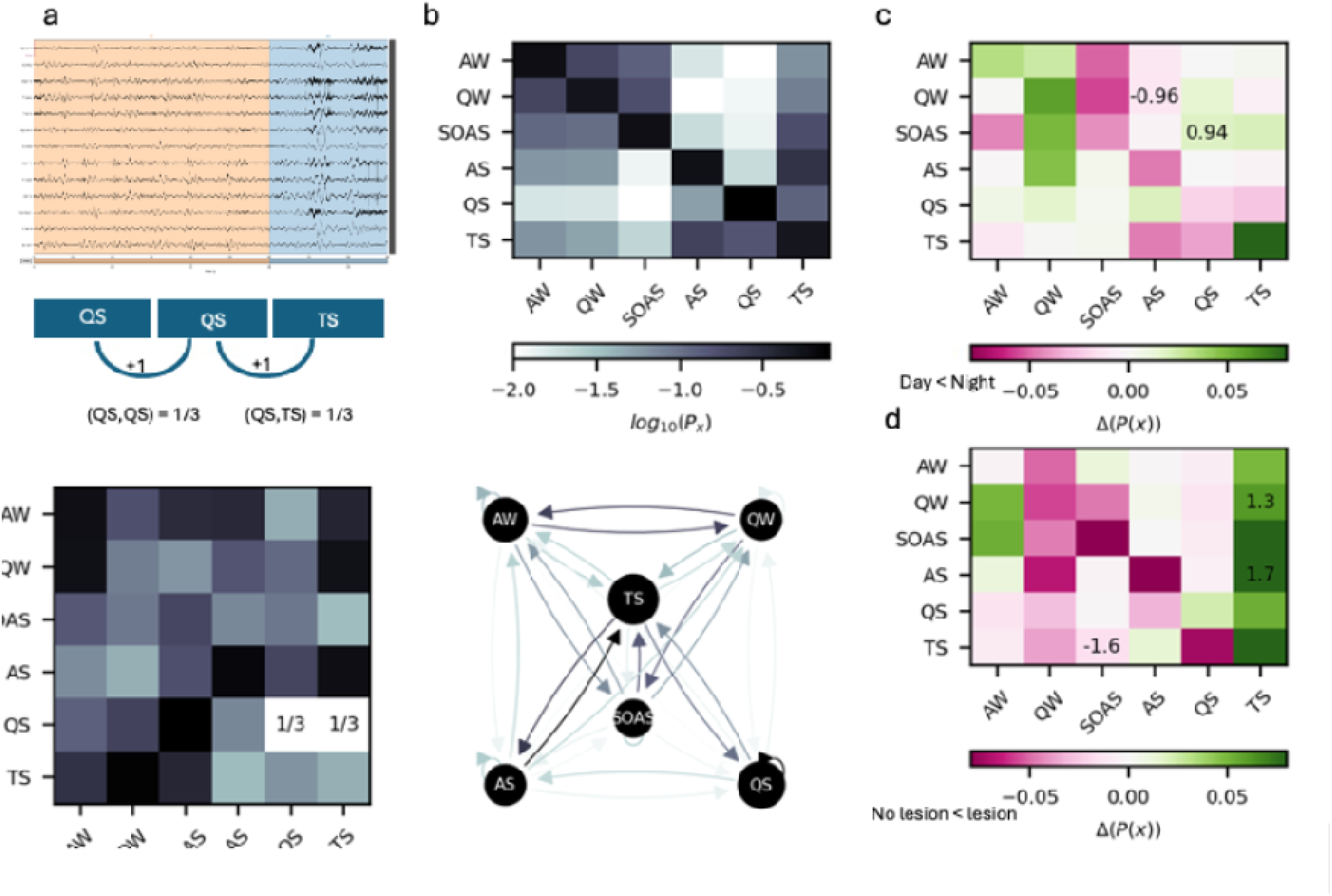
Sleep architecture of preterm infants and state-dependent transition probabilities. a) Algorithm for transition probabilities calculation. From sleep staging, we constructed a time-series of events where each sample represents a 30 seconds time window with a defined stage. Transition probabilities between two states are computed as the number of transitions over the total number of transitions. b) Transition probability matrix (top) and resulting flow graph. Probabilities are reported as log10(P) to increase contrast between states. Node size is proportional to state probability. Edge color is proportional to transition probability. c) Differences in transition probabilities for day and night (top) and non-lesion vs lesion (bottom) groups. Numbers represent Cohen’s d effect sizes for the significant tests only (p<0.05, Man-whitney U tests). AS, Active Sleep; QS, Quiet Sleep; SOAS, Sleep Onset Active Sleep; TS Transitional Sleep.

Furthermore, transition dynamics were affected by the presence of lesion in the MRI. Patients with lesions (major and minor) showed significantly higher transition probability from TS to SOAS, while patients with no lesions have significantly higher probability in transitioning to TS from both QW and AS (*Figure 3d*).

### Sleep states correlate with neurobehavioral state at 32 weeks PMA and at TEA

In the examined sample, a strong correlation of sleep states was seen with neurological, neurobehavioral and visual skills at 32 weeks of PMA (*Table* 2). Higher total sleep times were associated with HNNE scores and the NBAS at all domains. Quiet sleep and TS showed strong positive correlations, while SOAS and AS had a strong negative correlation to the outcomes assessed.

**Table 2.** Correlation among sleep distributions and neurobehavioral assessments performed at 32 weeks PMA. Correlation at the Spearman test * significance at the 0.05 level (2-tailed); ** significance at the 0.01 level (2-tailed). AS stands for Active Sleep, QS stands for Quiet Sleep; SOAS stands for Sleep Onset Active Sleep; TS stands for Transitional Sleep. Note that lower scores at visual assessment and higher scores at HNNE and NBAS correspond to better performances.

| $\rho$ | Total<br>Sleep time | AS+SOAS | SOAS | AS | QS | TS |
| --- | --- | --- | --- | --- | --- | --- |
| **HNNE total score** | **0.681\*** | -0.527 | **-0.707\*** | -0.359 | 0.563 | **0.651\*** |
| **NBAS** |  |  |  |  |  |  |
| Habituation | **0.718\*** | **-0.635\*** | -0.431 | **-0.707\*** | 0.216 | **0.695\*** |
| Orientation | **0.852\*** | **-0.755\*** | -0.671* | **-0.731\*** | **0.743\*** | 0.503 |
| State Organization | **0.871\*** | **-0.732\*** | -0.659* | **-0.732\*** | **.878\*\*** | 0.415 |
| State Regulation | **0.643\*** | **-0.731\*** | -0.455 | **-0.802\*\*** | 0.311 | **0.719\*** |
| Autonomic Organization | **0.845\*\*** | **-0.647\*** | -0.335 | **-0.766\*** | 0.467 | 0.591 |
| Motor Organization | **0.892\*** | **-0.695\*** | **-0.743\*** | **-0.647\*** | **0.850\*\*** | 0.419 |
| Reflexes | -0.321 | 0.595 | 0.574 | 0.383 | -0.431 | -0.547 |
| Supplementary Items | **0.683\*** | **-0.732\*** | -0.537 | **-0.683\*** | **0.823\*\*** | **0.927\*** |
| **Visual assessment score** | -0.125 | **-0.571\*** | **-0.682\*\*** | **-0.544\*** | -0.426 | -0.501* |

When looking at possible associations between amount of sleep during the night, the sample showed positive significant correlations (ρ > 0.8; *p* < 0.05) among total sleep time and TS with NBAS scales of Orientation and state organization, while AS and SOAS showed negative significant moderate (−0.6 < ρ < −0.8; *p* < 0.05) correlations with the same scale. Of noted difference, amount of AS+SOAS (together and separately) correlated with better visual performances. Observed associations at 32 weeks PMA, persisted with the same trend of significance, also at TEA (*Table 3*). Correlations among behavioural states transition probabilities and neurobehavioral assessments followed the same trend and are reported in the Supplementary Material (*Supplementary Tables 3-5*).

**Table 3.** Correlation among sleep distributions and neurobehavioral assessments performed at TEA. Correlation at the Spearman test * significance at the 0.05 level (2-tailed); ** significance at the 0.01 level (2-tailed). AS stands for Active Sleep, QS stands for Quiet Sleep; SOAS stands for Sleep Onset Active Sleep; TS stands for Transitional Sleep. Note that lower scores at visual assessment and higher scores at HNNE and NBAS correspond to better performances.

| $\rho$ | Total<br>Sleep time | AS+SOAS | SOAS | AS | QS | TS |
| --- | --- | --- | --- | --- | --- | --- |
| **HNNE total score** | **0.592\*** | -0.527 | **-0.647\*** | -0.348 | 0.663* | 0.563 |
| **NBAS** |  |  |  |  |  |  |
| Habituation | **0.695\*** | **-0.635\*** | -0.431 | **-0.507\*** | 0.216 | 0.216 |
| Orientation | **0.802\*** | **-0.705\*** | **-0.581\*** | **-0.689\*** | 0.654* | **0.543\*** |
| State Organization | **0.851\*** | **-0.732\*** | **-0.549\*** | **-0.682\*** | 0.789* | **0.578\*\*** |
| State Regulation | **0.842\*\*** | **-0.701\*** | -0.305 | 0.450 | **0.511\*** | 0.221 |
| Autonomic Organization | **0.878\*\*** | **-0.447\*** | -0.335 | -0.345 | 0.567 | 0.467 |
| Motor Organization | **0.791\*** | -0.395 | **-0.743\*** | -0.456 | **0.850\*\*** | **0.650\*** |
| Reflexes | -0.202 | 0.195 | 0.174 | 0.383 | -0.431 | -0.431 |
| Supplementary Items | 0.494 | -0.324 | -0.537 | -0.456 | **0.823\*\*** | **0.623\*** |
| **Visual assessment score** | -0.101 | **-0.631\*** | **-0.698\*\*** | **-0.545\*** | -0.526 | -0.426 |

**Table 4.** Correlation among sleep distributions and neurodevelopmental assessments performed at 6 months CA. Correlation at the Spearman test. No statistical significance was found. AS stands for Active Sleep, QS stands for Quiet Sleep; SOAS stands for Sleep Onset Active Sleep; TS stands for Transitional Sleep.

| $\rho$ | sleep stability | sleep stability | sleep stability | sleep stability | sleep stability | sleep stability |
| --- | --- | --- | --- | --- | --- | --- |
| Griffiths Mental Development<br>Scales – III | AW | QW | SOAS | AS | QS | TS |
| **Total Developmental Quotient** | 0,042 | 0,079 | -0,442 | -0,479 | 0,285 | 0,564 |
| **Learning foundations** | 0,012 | 0,287 | -0,304 | -0,322 | 0,219 | 0,474 |
| **Language and Communication** | 0,280 | -0,226 | -0,310 | -0,140 | 0,024 | 0,407 |
| **Eye and Hand Coordination** | -0,237 | 0,287 | -0,511 | -0,535 | 0,438 | 0,517 |
| **Personal-Social-Emotional** | 0,322 | -0,085 | -0,292 | -0,207 | 0,061 | 0,602 |
| **Gross Motor Skills** | -0,024 | -0,372 | -0,347 | -0,450 | 0,140 | 0,365 |

### Observed sleep states distributions have an impact on neurodevelopment at 6 months CA

At 6 months of CA, an influence of sleep states organization on total neurodevelopment, learning and fine motor skills was still visible (*Table 3*). Higher total sleep times were associated with better global developmental performances (Total Developmental Quotient), better learning foundation skills, language, eye-hand coordination, and social performances. Parallelly, amount of QS and AS was positively correlated with developmental outcome at 6 months of CA, while amount of SOAS was negatively correlated.

### Brain lesions impair sleep quantity and sleep distribution

Infants carrying brain lesions showed lower QS time (*p* = 0.008) accompanied by higher prevalence of total amount of AS (*p* = 0.003) and AS+SOAS (*p* = 0.007). A trend of shorter duration of both Total Sleep time and TS was observed (*Figure 4*). Of interest, VPI carrying brain lesions during night time spent less sleep time in QS (*p* = 0.009), and more time in total AS (AS+SOAS) and AS (*p* = 0.047 and *p* = 0.009 respectively) (*Figure 5*).

**Figure 4.**
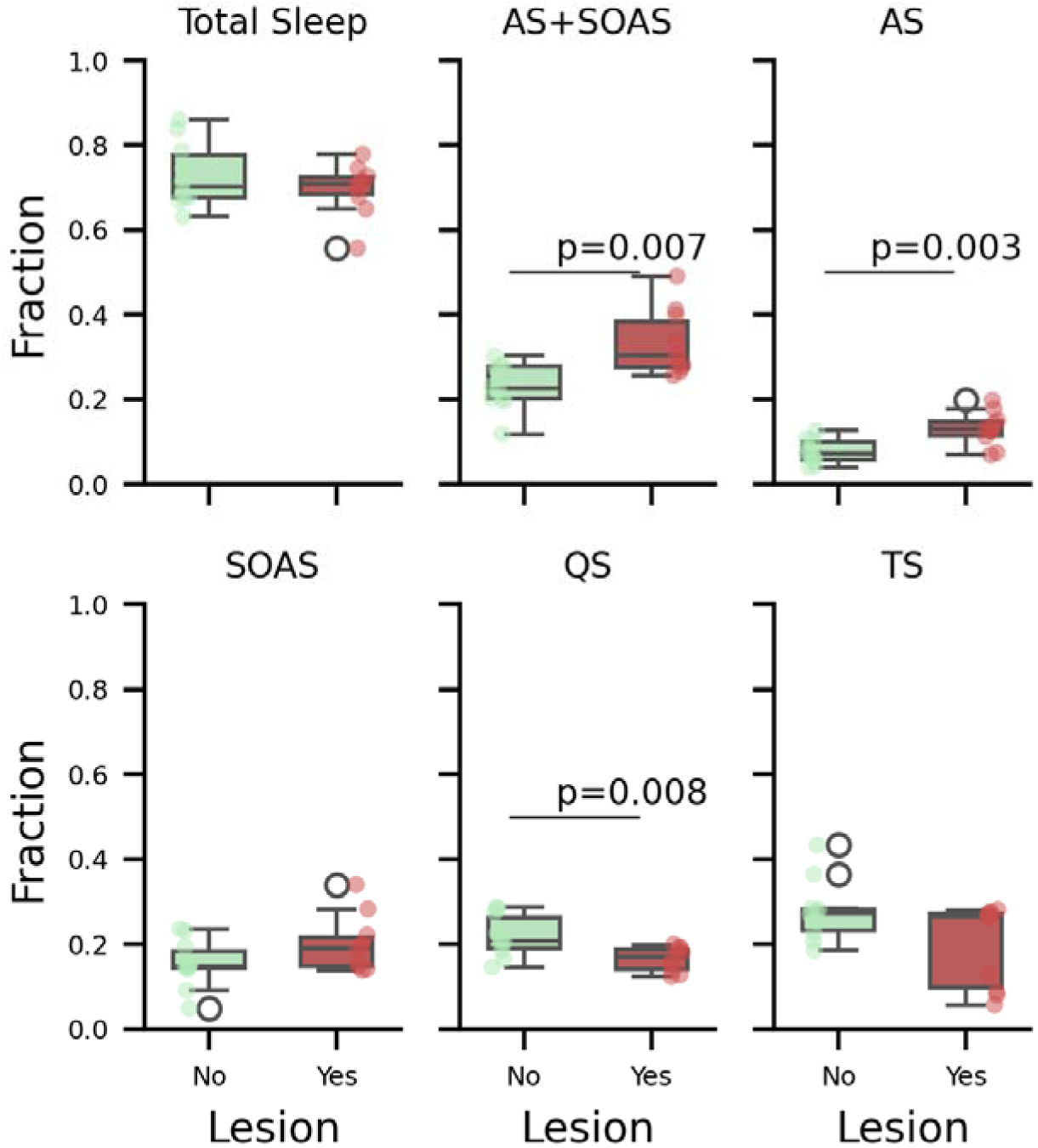
Average fraction of sleep events out of the total duration of sleep separated for patients without and with lesions. Figure 4. Average fraction of sleep events out of the total duration of sleep separated for patients without (green) and with lesions (red) visible in the MRI. Boxes present median (black line) and inter-quartile (black whiskers). Statistical significance is shown above individual tests with the relative pvalues. AS, Active Sleep; QS, Quiet Sleep; SOAS, Sleep Onset Active Sleep; TS Transitional Sleep.

**Figure 5.**
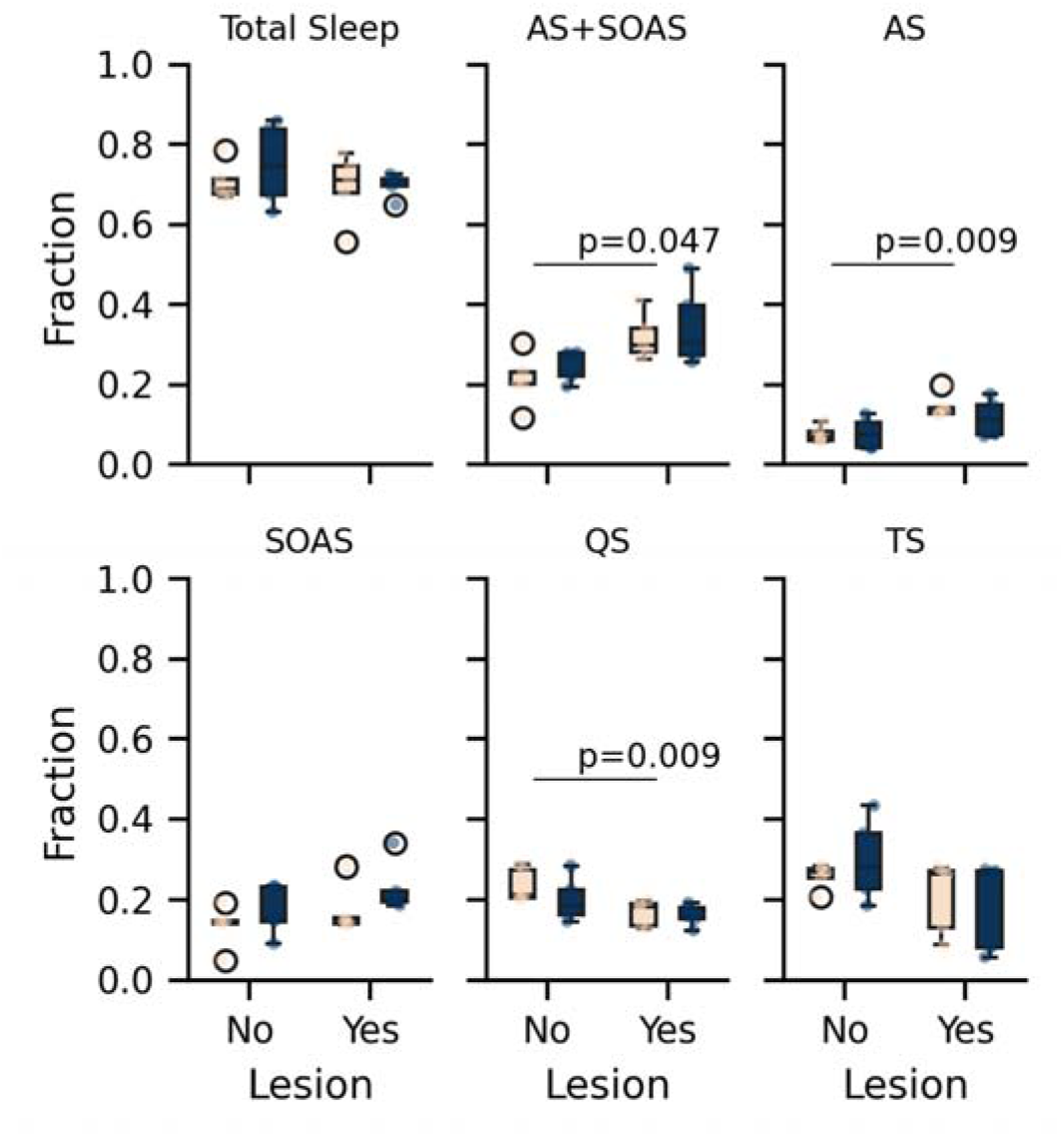
Average fraction of sleep events out of the total duration of sleep separated for day time and night time. Average fraction of sleep events out of the total duration of sleep separated Day (light orange) and Night (dark blue) as a function of lesions. Boxes present median (black line) and inter-quartile (black whiskers). Statistical significance is shown above individual tests with the relative pvalues. AS, Active Sleep; QS, Quiet Sleep; SOAS, Sleep Onset Active Sleep; TS Transitional Sleep.

### Severe-moderate lesions are associated to posterior slow-waves activity during SOAS in the left hemisphere

Spectral analyses revealed that VPI carrying moderate-severe brain lesions showed positive gradient of slow waves (slow δ and δ) in the left hemisphere during SOAS (*p =* 0.009 and *p =* 0.008 respectively). No other significant results were found. *Figure 6* illustrates gradients powers for SOAS in the left hemisphere.

**Figure 6.**
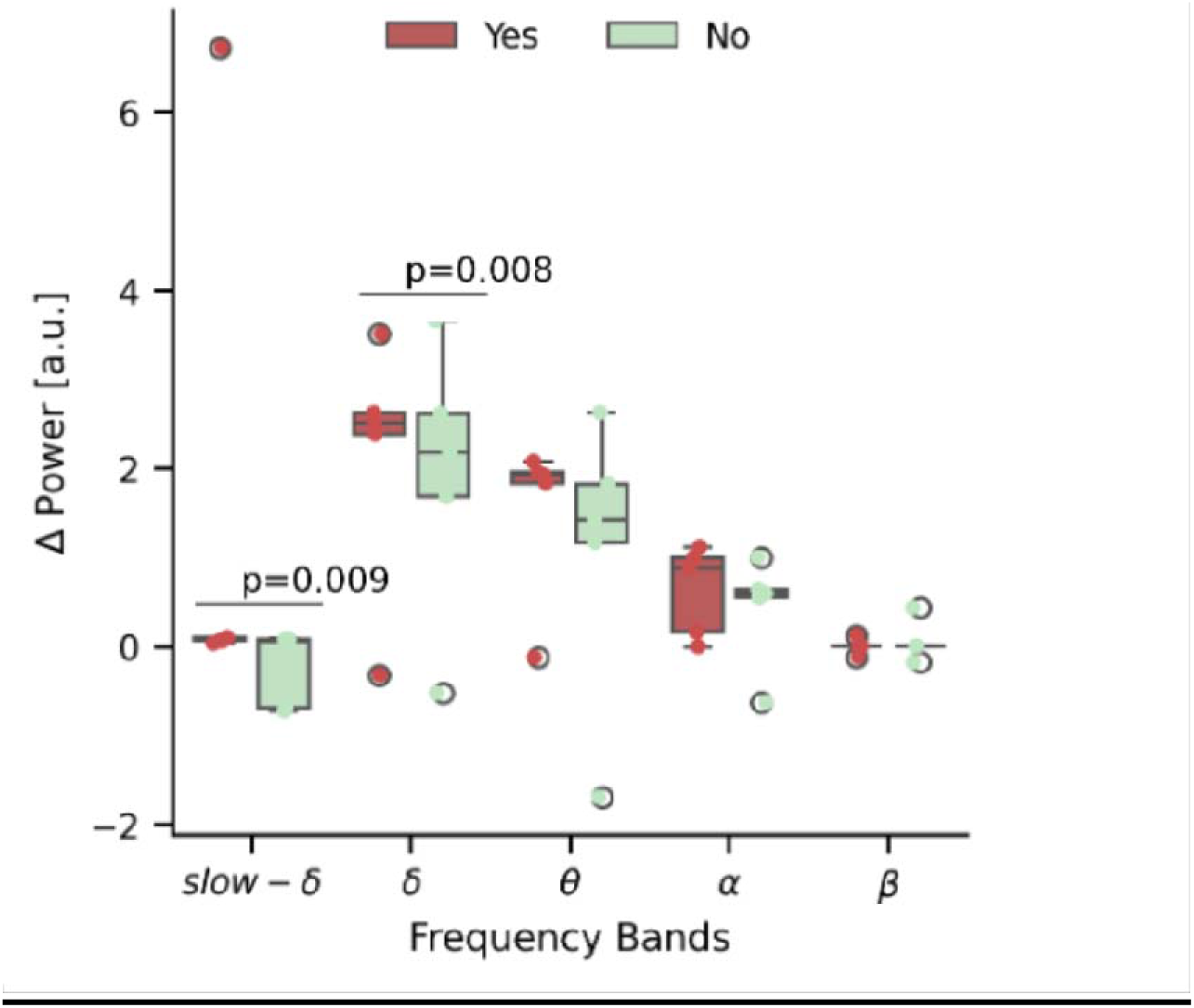
Distributions Δ spectra in the left hemisphere during Sleep Onset Active Sleep divided for VPI carrying or not severe-moderate brain lesions. Spectral power Δ was calculated starting from mean spectra over total spectral power. Posterior and anterior derivations were then confronted. *p* calculated with Mann-U Whitney test.

## Discussion

This pilot exploratory represents an initial step toward advancing analyses aimed at understanding the intrinsic meaning of sleep behavioral states in very preterm infants (VPI).

In the examined sample, our preliminary observations from 24-hour videoPSG recordings reveiled that the total amount of sleep, of Quiet Sleep (QS), and of Transitional Sleep (TS) significantly correlated with neurobehavioral assessments at 32 weeks of postmenstrual age (PMA), at term equivalent age (TEA), and with neurodevelopment at 6 months of corrected age (CA) in a sample of 10 VPI (*Tables 2-4*). Infants carrying brain lesions exhibited lower amount of QS and increased total Active Sleep (AS) time (which includes both Sleep Onset Active Aleep, SOAS, and AS). They also demonstrated a general trend of reduced duration of Total Sleep Time and TS (*Figures 4*), alongside a higher prevalence of a gradient for higher power of posterior slow activity (slow δ and δ) during SOAS in the left hemisphere’s posterior regions. Additionally, VPI carrying brain lesions during spent less night time sleep in QS and more time in total AS (AS+SOAS) and AS (*Figure 5*). It was also possible observing a trend of different sleep distribution across the day, with a prevalence of sleep (compared to wakefulness) in the night-time hours (20:00-08:00) in all types of states examined, with statistical significance for sleep onset active sleep (SOAS) (*Figure 2* and *Supplementary* Figure 1).

Sleep is a fascinating window that allows both understanding of physiological mechanisms of neuronal processes and pathological bases of the diseases, thus allowing target strategies of intervention. The importance sleep in participating in developmental processes is largely exemplified by the large amount spent in sleeping behaviours by foetuses, newborns and infants (Roffwarg et al., 1966).

Both NREM (Non-Rapid Eye Movement) and REM sleep have been linked to processes of neuronal plasticity and therefore cognition, memory reconsolidation, and emotional regulation (de Vivo et al., 2017; Li et al., 2017). Sleep loss can be detrimental, and has been linked to a systemic proinflammatory state and to brain microglial activation in absence of other neuroinflammatory processes (Bellesi et al., 2018), which, at early ages, may be the base for neurodevelopmental disorders.

Premature birth is a source of sleep perturbation in terms of functional connectivity and this is related to neurodevelopmental outcome later in childhood (Tokariev et al., 2019).

Although scientific evidence is emerging regarding sleep impairment among very preterm infants (Bennet et al., 2018; Dereymaeker et al., 2017; Gogou et al., 2019; Uchitel et al., 2022), the role of preterms’ sleep architecture in determining their well-known adverse neurological-neurobehavioral features and neurodevelopmental outcome remains largely unexplored. Analysis of behavioural observation or PSG recording during the 24 hours, described sleep distribution in smaller samples of preterm infants, and no correlation with neurological state and neurobehavioral outcome was provided (Biagioni et al., 2005; Curzi-Dascalova et al., 1993). Our prospective and ongoing study aimed to fill this gap.

Results of our preliminary observations show that total sleep time was a constant metrics of positive correlation with neurological, neurobehavioral and neurodevelopmental states. Reduced total sleep time is indeed a common finding among children suffering from neurodevelopmental disorders such as Autism Spectrum Disorders (Morgan et al., 2020), who also can present NREM sleep slow wave activity alterations, disorders in initiating and mantaining sleep (Miano et al., 2007). Total sleep time has been also designated as a predictor of late-in-childhood developmental outcome for critically ill newborns (Shellhaas et al., 2017). Sleep could become a feasible screening tool for very preterm infants’ wellbeing and future neurodevelopment.

From this set of data, active sleep at the start of sleep time (SOAS) and active sleep after quiet sleep (AS) were associated to more immature performances at neurological and neurobehavioral assessment while quiet sleep (QS) and transitional sleep (TS) correlated with more advanced performances at 32 weeks PMA and TEA, and also at 6 months of CA.

It can be postulated that in very preterm infants active sleep, especially in the form of SOAS, may be related to maturational processes that are likely to disappear during foetal and neonatal life, thus representing a marker of immaturity if present at this stage of development.

Active sleep, which equals to the activity of deep brain structures, is related to neuronal plasticity and to development of the sensory-motor processes, that are pillar to further cognition development, and follows, in animal models, fixed of deployment (Blumberg et al., 2020, 2022; Del Rio-Bermudez & Blumberg, 2018). Thus, it might be that active sleep, especially SOAS, may be a marker of immaturity of brain processes in very preterm infants.

On the other hand, quiet sleep has been instead related to long-range cortical connectivity in preterm infants at term of equivalent age (Tokariev et al., 2019) brain maturation, and postnatal cerebral maturation (Cailleau et al., 2020), and to memory consolidation later in life (Friedrich et al., 2020), being thus a possible marker for better brain organization if assessed early in very preterm infants.

In this context, the role for transitional sleep, that might mark the transition to a “more mature” state of brain organization, should be clarified. In fact, sparse literature postulated the maturational role played by transitional sleep among very preterm infants (de Weerd & van den Bossche, 2003).

It was also possible observing the clinical early proof that active sleep (both at the start of the sleep time, and after QS) may be associated to early visual performances at 32 weeks PMA and at TEA. Literature has already postulate the structural and functional link between visual cortex and AS/REM (Tokariev et al., 2019), to the point of saying that REM sleep can be considered “ocular twitches” through witch AS reorganise brain visual networks (Blumberg et al., 2022).

Of note, the Eye-Hand coordination domain of Griffiths at 6 months CA correlated negatively with amount of active sleep. This could be related to the fact that the scale assesses more complex brain functioning, which involves not only vision but also fine motor coordination and learning abilities, which are related to cortical brain areas, brainstem and cerebellum (Miall et al., 2001; Rizzo et al., 2020).

Moreover, infants carrying brain lesions (Ramenghi, 2015) showed lower Total Sleep time, higher prevalence of SOAS, and shorter duration of both QS and TS. The increment of SOAS+AS, although surprising, may be attribute to the ontogenetic role that it plays across development. AS/REM sleep is pillar for pruning and scaling synapses, acquisition of sensory-motor, visual, and adaptive behavioural skills. It might be postulated that exactly SOAS might be the designated to the process of post-injury synaptic repair and remodelling (Blumberg et al., 2020, 2022) after IVH. However, the significance of active sleep at sleep onset has to be clarified, as well as the appropriateness of using the term SOAS to distinguish it from active sleep after QS.

Indeed, at a preliminary quantitative analysis, they also showed a gradient for higher power of posterior slow activity during SOAS from the posterior cerebral regions in the left hemisphere (where the lesions were predominantly located). These observations may be also related to the pathophysiology of germinal matrix-intraventricular haemorrhages, that are associated with microstructural alterations caused by inflammatory processes starting from the blood remaining on the ependymal surface (Ballabh, 2014) located diffusely in the white matter, but mostly posteriorly (Tortora et al., 2018).

In the examined sample, we also observed different trend in sleep distributions across the 24 hours, with sleep time being more present during night hours. Although it is possible that nursing and medical procedures are likely to be performed more during day time thus disturbing preterm neonates’ sleep, presence of circadian rhythms among newborns and very preterm infants is debated since cortisol secretion seems to show a two-phase pattern (Spangler, 1991) and melatonin production starts from the 6th week of life, but is delayed in premature infants (Kennaway et al., 1992). Of interest, earlier melatonin production start is associate with better neurodevelopment (Tauman et al., 2002).

Biagioni et al, showed interesting clues regarding the distribution of sleep patterns and yawns across 24 hours. They assessed 11 neonates (of them 8 preterm infants, 5 early and 3 late preterms referred having a normal neurological outcome) and observed, from the electroencephalographic traces, a different distribution of sleep patterns among day and night (with a higher prevalence for the equivalent of QS during the day.

Although the limited sample size, this pilot study has to be considered representative of other larger published casistics, in terms of prevalence of categories of brain lesions (Malova et al., 2023; Uccella et al., 2023), trends of neurological and neurobehavioral performances (Canals et al., 2011; Mercuri et al., 2003; Ricci et al., 2010), and distribution of sleep behavioural states (André et al., 2010; Curzi-Dascalova et al., 1993, 1993).

To the best of the present knowledge, this was the first study investigating the short and mid-term outcome of sleep pattern among VPI. However, these data are only preliminary observations, where the small sample size did not allow for analyses of possible interrelationships of other prenatal/neonatal variables on Sleep patterns of VPI. They need to be furtherly corroborated by larger datasets and more accurate and complex metrics.

## Conclusions

Although the limited sample size and the exploratory nature of this study, data clearly show that very preterm infants present impaired sleep distribution across the 24 hours that is linked to neurological neurobehavioral set at early stages and at mid-term. The presence of brain lesions worsen this condition.

In this context, more accurate and measurable sleep care interventions should be designed and tested as possible protective tools of very preterm infants’ neurodevelopment.

## Data Availability

All data produced in the present study are available upon reasonable request to the authors

## Data availability

Raw data supporting the conclusions of this manuscript will be made available by the authors, without undue reservation, to any qualified researcher.

## Author contributions

SU and LN designed the study, SU constructed the databases, interpreted the data, made the statistical analysis, and drafted the manuscript. VM and MB helped in data collection and revised the manuscript. LN, VM, GB, GA, and LAR interpreted the data and revised the manuscript. All the authors discussed the results, commented on the manuscript, and approved the final draft.

## Acknowledgments.

The authors would like to acknowledge all the families that participated in the study, the nurses and technical personnel working at the Neonatology Unit, and at the Child Neuropsychiatry Unit of the IRCCS Gaslini Children’s Hospital, and the association EuBrain, which intellectually and practically spent time in understanding the nature and the problem of the developing brain of premature infants.

## Institutional Review board statement

The local ethics committee (Comitato Etico Territoriale, Genoa, Italy) approved this study, and the participating parents provided online informed consent in accordance with the Declaration of Helsinki.

## Patients consent statement

Parents’ of the enrolled subjects have received appropriate information and signed informed consent.

