## Supplementary material for "Sleep architecture correlates with neurological and neurobehavioral short- and mid-term outcome in a sample of very preterm infants"

*Supplementary Table 1.* Prenatal, intrapartum, and neonatal features of the investigated sample

|  | **Total (n 10)** |
| --- | --- |
| Multiple gestation, *n (%)* | 4 (40%) |
| Monochorial twins, *n* (%) | 3 (30%) |
| IUGR, *n (%)* | 5 (50%) |
| Maternal hypertension, *n (%)* | 3 (30%) |
| Gestational diabetes, *n (%)* | 1 (10%) |
| Absent/incomplete antenatal steroid course, *n (%)* | 2 (20%) |
| pPROM, *n (%)* | 1 (10%) |
| Birth by caesarean section, *n (%)* | 7 (70%) |
| Gestational age (weeks), *mean (sds)* | 30.5 (3.2) |
| Birth weight (grams), *mean (sds)* | 1300.0 (344.7) |
| Apgar at 1^st^ minute, *mean (sds)* | 6.4 (2.1) |
| Apgar at 5^th^ minute, *mean (sds)* | 8.0 (1.3) |
| Intubation during first 72 h, *n (%)* | 4 (40%) |
| Pneumothorax during first 72 h, *n (%)* | 2 (20%) |
| HFO during first 72 h, *n (%)* | 0 (0%) |
| MV > 14 days, *n (%)* | 1 (10%) |
| Multiple surfactant doses, *n (%)* | 2 (20%) |
| EOS, *n (%)* | 1 (10%) |
| LOS, *n (%)* | 1 (10%) |
| NEC, *n (%)* | 1 (10%) |
| Surgery, *n (%)* | 1 (10%) |
| BPD, *n (%)* | 2 (20%) |
| ROP (stage III-IV), *n (%)* | 1 (10%) |
| Mild brain lesions | 3 (30%) |
| Moderate/severe brain lesions | 2 (20%) |

*Legend.* EOS, early-onset sepsis; GA, gestational age; IUGR, intrauterine growth restriction; HFO, high-frequency oscillations; MV, mechanical ventilation; NEC, necrotizing enterocolitis; EOS, early-onset sepsis; LOS, late-onset sepsis; pPROM, premature prolonged rupture of membranes; ROP, retinopathy of prematurity

*Supplementary Table* *2*. Longitudinal Neurobehavioral assessments of the studied sample

| **Evaluation at 32 weeks PMA** | *median (min-max)* | *n (%) normal scores* |
| --- | --- | --- |
| HNNE total score | 28 (1.50-30) |  |
| NBAS assessment  Habituation  Orientation  State Organization  State Regulation  Autonomic Organization  Motor Organization  Reflexes  NBAS Supplementary Items | 6.8 (4.3-8.0)  7.4 (2.8-8)  4.6 (3.3-6.3)  6.6 (3.8-7.5)  5.2 (3.7-6.3)  5.1 (4.4-6.5)  4.0 (1-15)  7.1 (4.6-7.4) |  |
| Visual assessment score | 3.5 (2-11) |  |
| **Evaluation at TEA total score** *median (min-max)* |  |  |
| HNNE total score | 32.0 (13.5-33.5) |  |
| NBAS assessment  Habituation  Orientation  State Organization  State Regulation  Autonomic Organization  Motor Organization  Reflexes  NBAS Supplementary Items | 7.3(4.3-7.8)  7.8 (3.3-8.1)  5.5 (3.3-7.8)  6.5 (4.3-8.0)  7.0 (4.0-8.0)  6.8 (4.4-7.5)  0.5 (0-13)  7.5 (4.6-8.3) |  |
| Visual assessment score | 1.5 (0-8) |  |
| **Evaluation at 6 months CA** |  |  |
| Griffiths Mental Development Scales III  Total Developmental Quotient  Learning foundations  Language and Communication  Eye and Hand Coordination  Personal-Social–Emotional  Gross Motor Skills | 117.5 (55-128)  126.5 (56-138)  105 (40-120)  110 (61-133)  114 (67-128)  113.5 (54-118) |  |

*Legend.* HNNE, Hammersmith Neonatal Neurological Examination; NBAS, Newborn Behavioral Assessment Scale. Note that lower scores at visual assessment and at NBAS reflex scale and higher scores at HNNE, NBAS and Griffiths-III correspond to better performances.

*Supplementary Figure 1.* Fraction of sleep compared to wake periods across the entire 24h recoding.

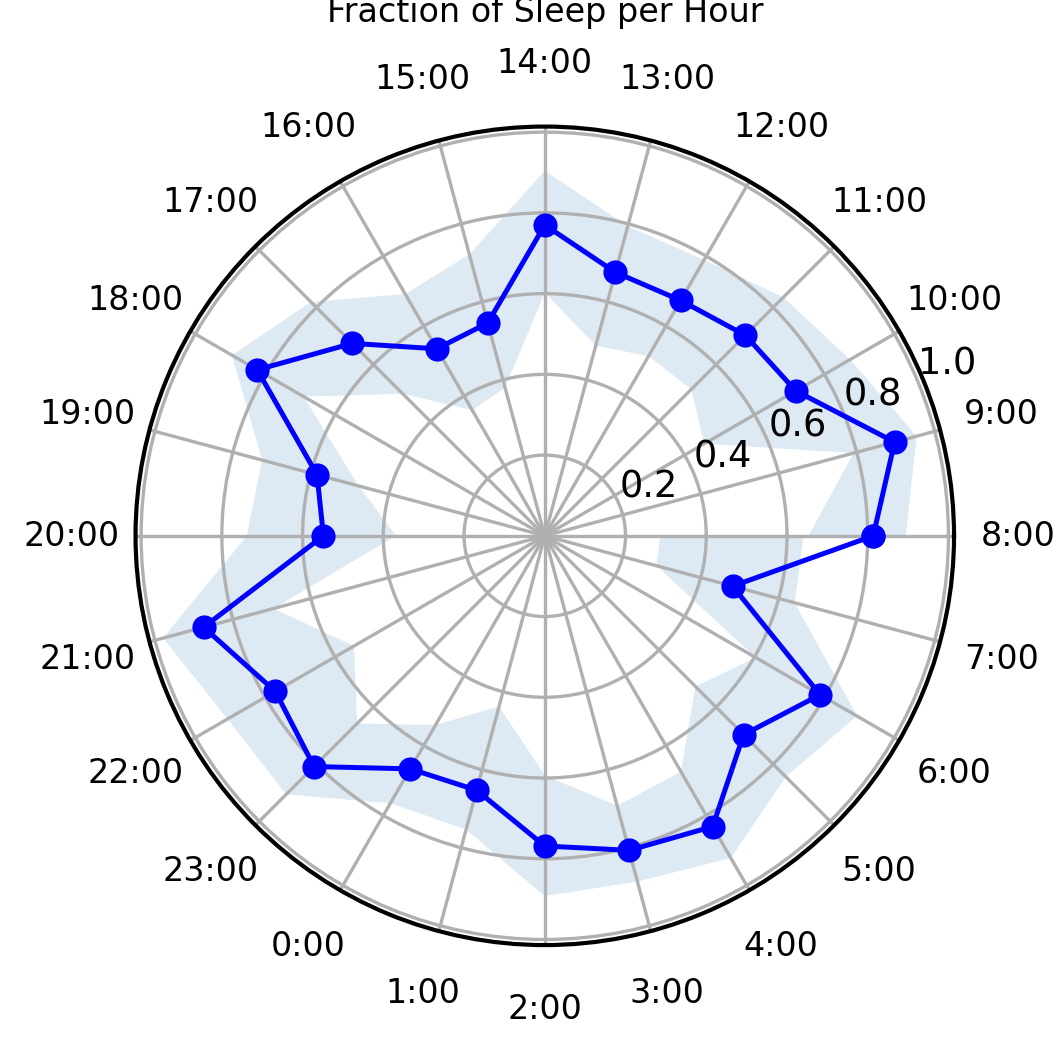

***Legend.*** Data are shown in circular plot over the 24h long day. Coloured arcs represent the separation between day (8:00-> 20:00) and night (20:00->8:00). Thick line represents the mean across the cohort. Shaded area represents the confidence intervals around the mean (bootstrap CI: 5% two tails, N=9999 resampling with replacement)

| *ρ* | **sleep stability AW** | **sleep stability QW** | **sleep stability SOAS** | **sleep stability AS** | **sleep stability QS** | **sleep stability TS** |
| --- | --- | --- | --- | --- | --- | --- |
| **HNNE total score** | -0,232 | -0,021 | -0,585 | **-0,750*** | -0,604 | 0,329 |
| **NBAS** |  |  |  |  |  |  |
| Habituation | -0,049 | 0,119 | -0,555 | -0,543 | -0,604 | 0,335 |
| Orientation | 0,091 | 0,006 | -0,394 | -0,515 | -0,261 | 0,600 |
| State Organization | 0,239 | 0,111 | -0,374 | -0,423 | -0,313 | 0,607 |
| State Regulation | -0,172 | 0,034 | -0,646 | **-0,702**** | -0,375 | 0,529 |
| Autonomic Organization | -0,322 | 0,454 | -0,602 | **-0,754**** | **-0,669**** | 0,395 |
| Motor Organization | 0,283 | 0,012 | -0,295 | -0,258 | -0,178 | 0,394 |
| Reflexes | -0,313 | 0,172 | **-0,742*** | **-0,841**** | -0,583 | 0,436 |
| Supplementary Items | -0,031 | 0,006 | 0,306 | 0,514 | 0,563 | -0,183 |
| **Visual assessment score** | -0,045 | 0,065 | 0,370 | 0,324 | 0,311 | -0,461 |

*Supplementary Table* *3.* Correlation among transition probability and neurobehavioral assessments performed at 32 weeks postmenstrual age.

*Legend.* Correlation at the Spearman test * significance at the 0.05 level (2-tailed); ** significance at the 0.01 level (2-tailed).

AS stands for Active Sleep, QS stands for Quiet Sleep; SOAS stands for Sleep Onset Active Sleep; TS stands for Transitional Sleep. Note that lower scores at visual assessment and higher scores at HNNE and NBAS correspond to better performances.

*Supplementary Table* *4.* Correlation among transition probability and neurobehavioral assessments performed at Term Equivalent Age.

| *ρ* | **sleep stability AW** | **sleep stability QW** | **sleep stability SOAS** | **sleep stability AS** | **sleep stability QS** | **sleep stability TS** |
| --- | --- | --- | --- | --- | --- | --- |
| **HNNE total score** | **-0,707** | 0,413 | -0,463 | **-0,756** | -0,799 | -0,244 |
| **NBAS** |  |  |  |  |  |  |
| Habituation | -0,109 | 0,104 | **-0,790**** | **-0,663**** | -0,498 | **0,644*** |
| Orientation | -0,333 | 0,043 | **-0,721*** | **-0,758*** | -0,600 | 0,297 |
| State Organization | -0,155 | 0,168 | **-0,665*** | **-0,715*** | -0,404 | **0,690*** |
| State Regulation | -0,396 | 0,122 | -0,421 | **-0,726*** | -0,439 | 0,317 |
| Autonomic Organization | -0,127 | 0,030 | -0,515 | -0,612 | -0,236 | 0,624 |
| Motor Organization | -0,366 | -0,049 | -0,610 | **-0,665*** | -0,518 | 0,177 |
| Reflexes | -0,267 | -0,171 | -0,547 | **-0,705*** | -0,389 | 0,426 |
| Supplementary Items | 0,257 | -0,160 | **0,690**** | **0,816*** | **0,627*** | -0,452 |
| **Visual assessment score** | 0,175 | -0,072 | **0,681**** | 0,623 | 0,519 | -0,253 |

*Legend.* Correlation at the Spearman test * significance at the 0.05 level (2-tailed); ** significance at the 0.01 level (2-tailed).

AS stands for Active Sleep, QS stands for Quiet Sleep; SOAS stands for Sleep Onset Active Sleep; TS stands for Transitional Sleep. Note that lower scores at visual assessment and higher scores at HNNE and NBAS correspond to better performances.

*Supplementary Table* *5.* Correlation among transition probability and neurobehavioral assessments performed at 6 months of Corrected Age.

| *ρ*  **Griffiths Mental Development Scales – III** | **sleep stability AW** | **sleep stability QW** | **sleep stability SOAS** | **sleep stability AS** | **sleep stability QS** | **sleep stability TS** |
| --- | --- | --- | --- | --- | --- | --- |
| **Total Developmental Quotient** | **0.852^**^** | **-0.905^**^** | **-0.929^**^** | -0.299 | **0.881^**^** | **0.848^**^** |
| **Learning foundations** | **0.838^**^** | -0.587 | **-0.850^**^** | 0.06 | 0.771 | **0.886^**^** |
| **Language and Communication** | 0.591 | -0.431 | -0.599 | -0.199 | 0.707 | 0.299 |
| **Eye and Hand Coordination** | **0.747^*^** | -0.659 | -0.635 | -0.265 | 0.563 | **0.743^*^** |
| **Personal-Social–Emotional** | **0.695*** | -0.623 | -0.695 | -0.343 | **0.802^*^** | 0.347 |
| **Gross Motor Skills** | 0.338 | -0.619 | -0.667 | -0.18 | **0.833^*^** | 0.167 |

*Legend.* Correlation at the Spearman test * significance at the 0.05 level (2-tailed); ** significance at the 0.01 level (2-tailed).

AS stands for Active Sleep, QS stands for Quiet Sleep; SOAS stands for Sleep Onset Active Sleep; TS stands for Transitional Sleep.
